# Sociodemographic differences in patient experience with virtual care during COVID-19

**DOI:** 10.1101/2021.07.19.21260373

**Authors:** Payal Agarwal, Rick Wang, Christopher Meaney, Sakina Walji, Ali Damji, Navsheer Gill Toor, Gina Yip, Debbie Elman, Tiffiany Florindo, Susanna Fung, Melissa Witty, Thuy Nga Pham, Noor Ramji, Tara Kiran

## Abstract

**Purpose:** We sought to understand patients’ care-seeking behaviours during the pandemic, their use and views of different virtual care modalities, and whether these differed by sociodemographic factors.

**Methods:** We conducted a multi-site cross-sectional patient experience survey at thirteen academic primary care teaching practices between May and June of 2020. An anonymized link to an electronic survey was sent to a subset of patients with a valid email address on file; sampling was based on birth month. For each question, the proportion of respondents who selected each response was calculated, followed by a comparison by sociodemographic characteristics using chi-squared tests.

**Results:** In total, 7482 participants responded to the survey. Most received care from their primary care clinic during the pandemic (67.7%, 5068/7482), the majority via phone (82.5%, 4195/5086). Among those who received care, 30.53% (1509/4943) stated that they delayed seeking care because of the pandemic. Most participants reported a high degree of comfort with phone (92.4%, 3824/4139), video (95.2%, 238/250) and email or messaging (91.3%, 794/870). However, those reporting difficulty making ends meet, poor or fair health, and arriving in Canada in the last 10 years reported lower levels of comfort with virtual care and fewer wanted their practice to continue offering virtual options after the pandemic.

**Conclusions:** Our study suggest that newcomers, people living with a lower income, and those reporting poor or fair health have a stronger preference and comfort for in-person primary care. Further research should explore potential barriers to virtual care and how these could be addressed.

## Introduction

The COVID-19 pandemic has dramatically shifted the way health care is delivered and experienced by patients. Primary care practices in Canada, the US and elsewhere, rapidly switched to a virtual first approach in the first wave of the pandemic to limit transmission of the SARS-CoV2 virus and conserve Personal Protective Equipment^1^. A study from Ontario found that shortly after the pandemic was declared, in-office visits reduced by 79% and virtual care increased 56-fold, comprising 71% of primary care physician visits^2^. While this approach supported immediate public health goals, its impact on access, receipt of patient centered evidence-based care, and longer -term health outcomes is unclear^3^. As health systems consider what the “new normal” should look like, an examination of these impacts will be crucial.

This shift to a virtual-first approach in primary care has raised several concerns. Clinicians have noted anecdotally that patients with worrisome symptoms are delaying care^4^. Some note that the switch to virtual care may make care more accessible^5–8^, while others have highlighted barriers certain populations face in accessing virtual care^9–12^. Prior studies suggest patient characteristics including older age and lower income may limit one’s ability to benefit from digital health and virtual care services^13–14^. In addition, patients may not have access to required technologies such as a phone or internet access^15–16^. Despite this, very little literature to date is available on patient experience during the COVID-19 pandemic and how these differ by sociodemographic characteristics. Most existing studies on patient experience during COVID-19 are from acute care, and do not stratify experience based on patient demographics^17–21^.

We conducted a patient survey at multiple academic primary care clinics in Ontario, Canada to better understand patient experience during COVID-19. We were interested in patients’ care-seeking behaviours, their use and views of different virtual care modalities, and whether these differed by sociodemographic factors.

## Methods

### Study design and setting

We conducted a multi-site cross-sectional survey to understand patient experience during the COVID-19 pandemic at thirteen core teaching practices affiliated with the University of Toronto Department of Family and Community Medicine situated in the Greater Toronto Area, Canada’s largest metropolitan area. Participating practices were located in Toronto and surrounding areas including Mississauga, Markham, and Barrie. Practices range in size from roughly 11 physicians serving 14,000 patients to 80 physicians serving 50,000 patients; some have multiple locations. Physicians in all teaching practices are part of Family Health Organizations and formally enroll patients, have shared responsibility for after-hours care, and are paid primarily by age-sex adjusted capitation; twelve of the thirteen sites were part of Family Health Teams that included non-physician health professionals such as nurses, nurse practitioners, social workers and dieticians.

The survey was developed to directly inform quality improvement (QI) efforts at participating sites during COVID-19. The initiative was formally reviewed by institutional authorities at Unity Health Toronto and deemed to neither require Research Ethics Board approval nor written informed consent from participants.

### Study Population and Recruitment

A link to an open electronic patient experience survey is emailed every quarter to a subset of patients with a valid email address on file; sampling each quarter is based on birth month with all eligible patients receiving a survey in a given year. The current analysis focuses on patients who were sent the survey between May and June of 2020 because they had a birthday during the months of March, April or May of 2020. Each site distributed an anonymized link to patients in the manner by which they usually communicate electronically to patients (i.e. by email or using a secure messaging service). In some cases, the email address on file may belong to a family member or caregiver to allow them the option of filling out the survey on behalf of the patient.

Recruitment was done in English, with one site also doing recruitment and survey completion in French. No incentives were provided to participants.

### Survey Design

The survey was developed collaboratively by the family physicians who had a QI leadership role at participating sites. Where possible, questions were informed by existing surveys including the Commonwealth Fund International Health Policy Survey^22–23^ and the Ontario Primary Care Experience Survey, which was developed as part of a larger Primary Care Performance Measurement strategy to measure the performance across 9 domains^24–25^. The survey went through several iterations based on feedback from practice QI teams, a survey methodologist, a biostatistician, patient education and engagement specialists, and patient and family advisors. A paragraph at the start of the survey outlined the purpose of the survey, the reason they were being asked to participate, and highlighted that the survey was voluntary and anonymous. The final survey was prepared in Qualtrics software, a digital platform to capture experience data, and included 43 potential questions over 5 thematic domains including: 1) seeking and delaying care, 2) use and comfort with virtual care, 3) urgent care access, 4) patient centeredness and 5) patient demographic and contextual factors. Participants could end the survey at any point and were able to review previously answered questions before submission. (See appendix for full survey)

### Data Collection and Storage

Data collected via the electronic survey were stored on Qualtrics. All data was downloaded onto a secure research server at the University of Toronto. A script was run to remove any potentially identifying information including 1) IP addresses, 2) Email address, 3) longitude/latitude coordinates and 4) any free text fields (which may contain unstructured protected health information).

### Statistical Analysis

We performed an initial descriptive statistics analysis on the responses of all participants across all sites who answered at least one question in the survey. For each question, we calculated the proportion of respondents who selected each response. We then compared patient responses by sociodemographic characteristics including age, gender, education, self-reported financial issues, immigration status, primary language, self-reported health, and usual primary care provider (PCP). P values were calculated using chi-squared tests and all data analysis was conducted using R version 4.0.

## Results

The survey link was emailed to 32,307 patients at 13 practices (see Appendix). We presented sociodemographic data (Table 1) on the 7482 participants who answered one or more questions in the survey (23.3% response rate). Sixty-five percent of respondents were female (4379/6713) and 78.3% (5159/6588) reported having a college, university or graduate degree. Nine percent of respondents (590/6556) reported trouble making ends meet at the end of the month, while 29.0% (1928/6656) were not born in Canada (with 16.9% of these having arrived in the last 10 years). Eight percent (553/6851) reported filling out the survey on behalf of a family member. Fifty-five surveys were completed in French.

**Table 1.**
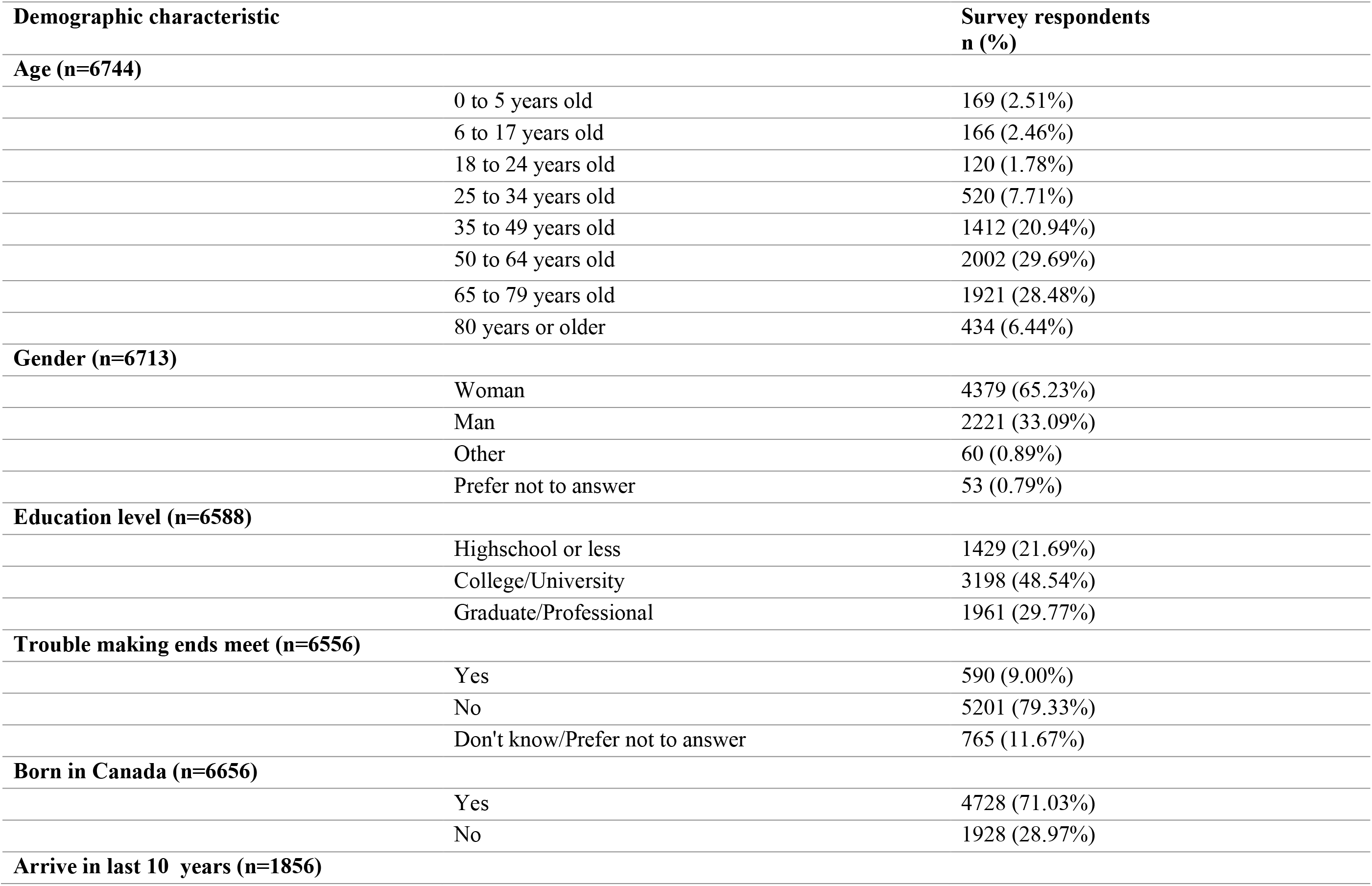
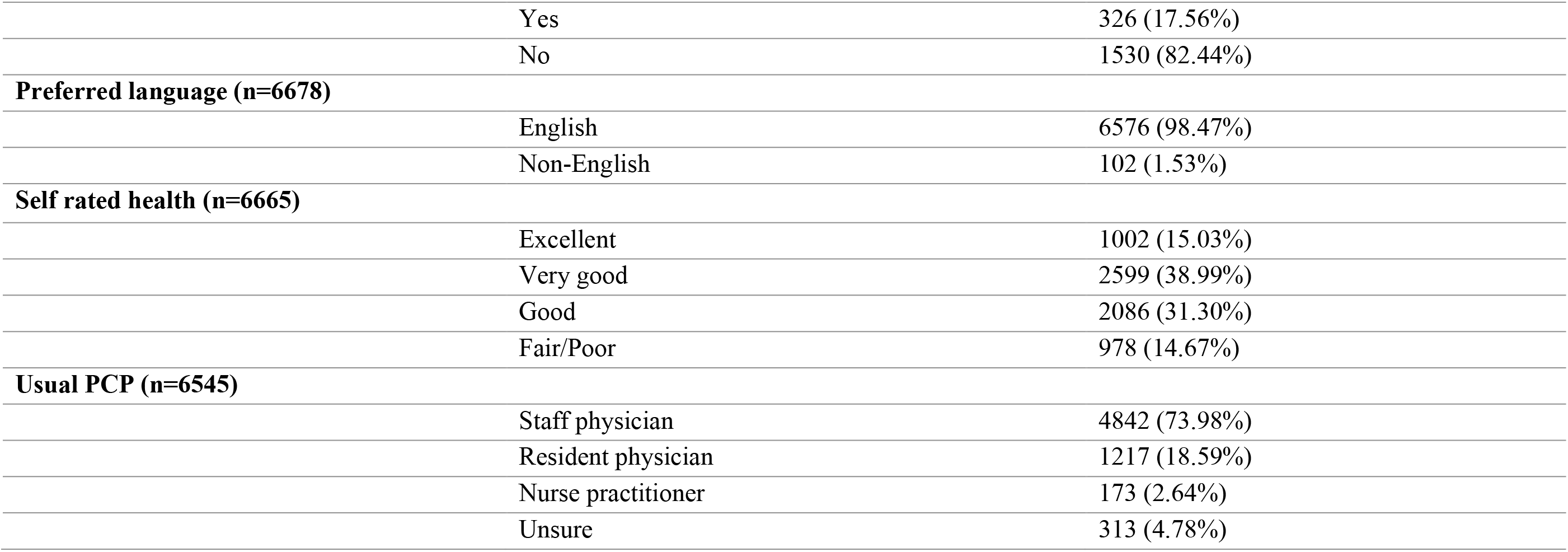
Demographic characteristics of survey respondents (n=7482)

### Care-seeking during COVID-19

Of all respondents, 67.7% (5068/7482) reported they received care in some way from their primary care clinic since the start of the pandemic. Financial status and self-reported health were significantly associated with care seeking behaviours during the pandemic (p<0.001); a higher proportion of patients who noted trouble making ends meet (yes: 74.8%, 441/590, no: 65.3%, 3396/5201, prefer not to answer: 68.0%, 520/765) and those with lower self-rated health (fair or poor: 76.9%, 752/978, excellent: 59.8%, 599/1002) reported receiving care during the study period. Of the 32.3% (2414/7482) of patients who did not receive care during the study period, the most commonly cited reasons were that patients had no health need (72.4%) and patients were worried about safety (9.5%).

Of the 5068 patients who reported receiving care at their primary care practice during the pandemic, 30.5% (1509/4943) stated that they delayed seeking care because of the pandemic.

Gender, age, education level, financial status and self-reported health status were significantly associated with differences in seeking care (p<0.05 for all); for example, a higher proportion of those with trouble making ends meet and those with lower self-rated health reported delays in seeking care (see Table 2).

**Table 2.** Proportion of respondents who received care at their primary care practice who reported that they delayed seeking care because of the pandemic, by sociodemographic characteristic.

| **Demographics** |  | **Delayed Care**<br>**n (%)** | **P-value** |
| --- | --- | --- | --- |
| **All** |  | 1509 (30.53%) |  |
| **Age** |  |  |  |
|  | <18 years old | 61 (29.90%) | <0.01 |
|  | 18-34 years old | 158 (35.03%) |  |
|  | 35-64 years old | 704 (31.44%) |  |
|  | 65+ years old | 429 (26.85%) |  |
| **Gender** |  |  |  |
|  | Woman | 932 (31.65%) | <0.001 |
|  | Man | 376 (26.07%) |  |
|  | Other | 20 (42.55%) |  |
|  | Prefer not to answer | 18 (46.15%) |  |
| **Education** |  |  |  |
|  | Highschool or less | 238 (24.74%) | <0.001 |
|  | College/University | 637 (30.09%) |  |
|  | Graduate/Professional | 443 (33.95%) |  |
| **Trouble making ends meet** |  |  |  |
|  | Yes | 188 (42.63%) | <0.001 |
|  | No | 928 (27.33%) |  |
|  | Don't know/Prefer not to answer | 196 (37.69%) |  |
| **Born in Canada** |  |  |  |
|  | Yes | 966 (30.92%) | 0.07 |
|  | No | 366 (28.09%) |  |
| **Arrive <10 years** |  |  |  |
|  | Yes | 66 (29.33%) | 0.79 |
|  | No | 289 (28.20%) |  |
| **Preferred Language** |  |  |  |
|  | English | 1322 (30.20%) | 0.49 |
|  | Non-English | 24 (34.78%) |  |
| **Self rated health** |  |  |  |
|  | Excellent | 153 (25.54%) | <0.001 |
|  | Very good | 475 (28.84%) |  |
|  | Good | 433 (30.20%) |  |
|  | Fair/Poor | 275 (36.57%) |  |
| **Usual PCP** |  |  |  |
|  | Staff physician | 976 (30.13%) | 0.47 |
|  | Resident physician | 246 (29.85%) |  |
|  | Nurse practitioner | 45 (36.59%) |  |
|  | Unsure | 52 (29.21%) |  |

### Use and perceptions of virtual care

Eighty-two (4195/5086) percent of participants reported receiving care by phone, 30.5% (1553/5086) in-person, 17.4% (886/5086) via email or secure messaging, and 5.1% (260/5086) via video. Age, immigration status and self-rated health were significantly associated with differences in receiving in person care (p<0.05 for all); the proportion who reported receiving in-person care was lower among those over the age of 65, those not born in Canada and those with lower self-rated health (Table 3). Women, young adults, those who rate their health as fair or poor and those who reported trouble making ends meet reported higher rates of phone use (see Table 3). Age and education level were significantly associated with differences in using email and secure messaging (p<0.001 for all); those over the age of 65 (16.9%, 269/1592) and those with a high school degree or less (13.5%, 130/962) reported less use of email and secure messaging relative to other groups.

**Table 3.**
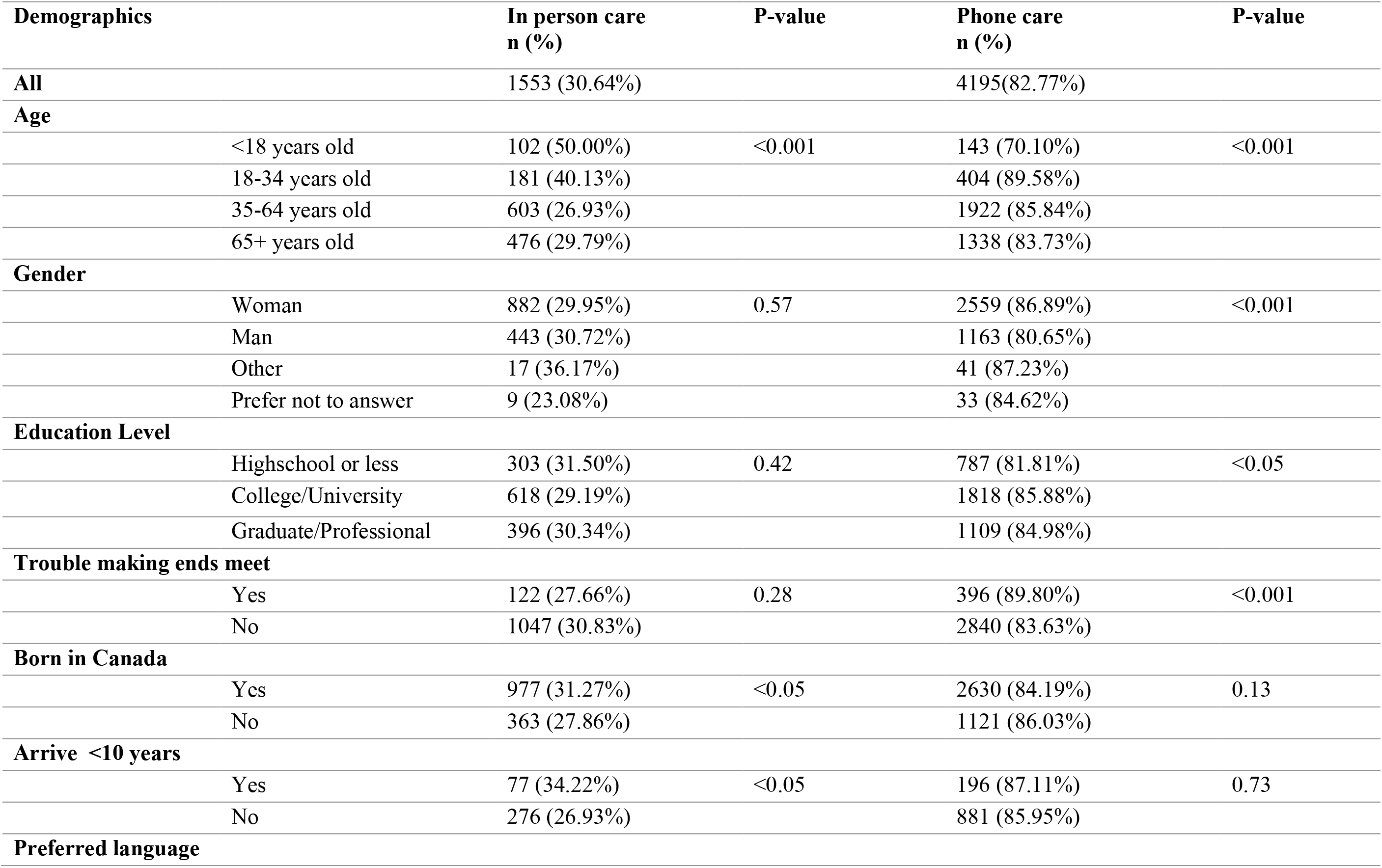
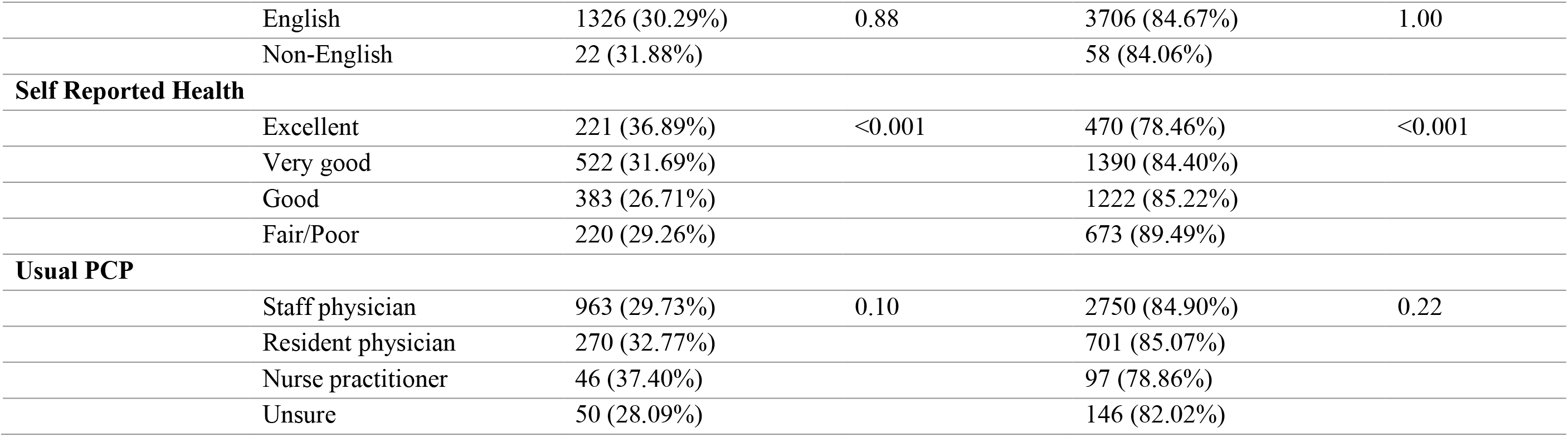
Percentage of patients who reported receiving care by phone and in person during the pandemic, by sociodemographic characteristic.

Overall, most respondents indicated they were extremely or somewhat comfortable with the privacy and security of virtual modalities including phone (92.4%, 3824/4139), video (95.2%, 238/250) and email or secure messaging (91.3%, 794/870). Financial status, immigration status and self-reported health status were significantly associated with differences in comfort with virtual care use (p<0.05 for all); those having trouble making ends meet, those not born in Canada and those rating their health as fair or poor reported lower levels of comfort with phone calls and email or secure messaging relative to other groups (see Table 4).

**Table 4.**
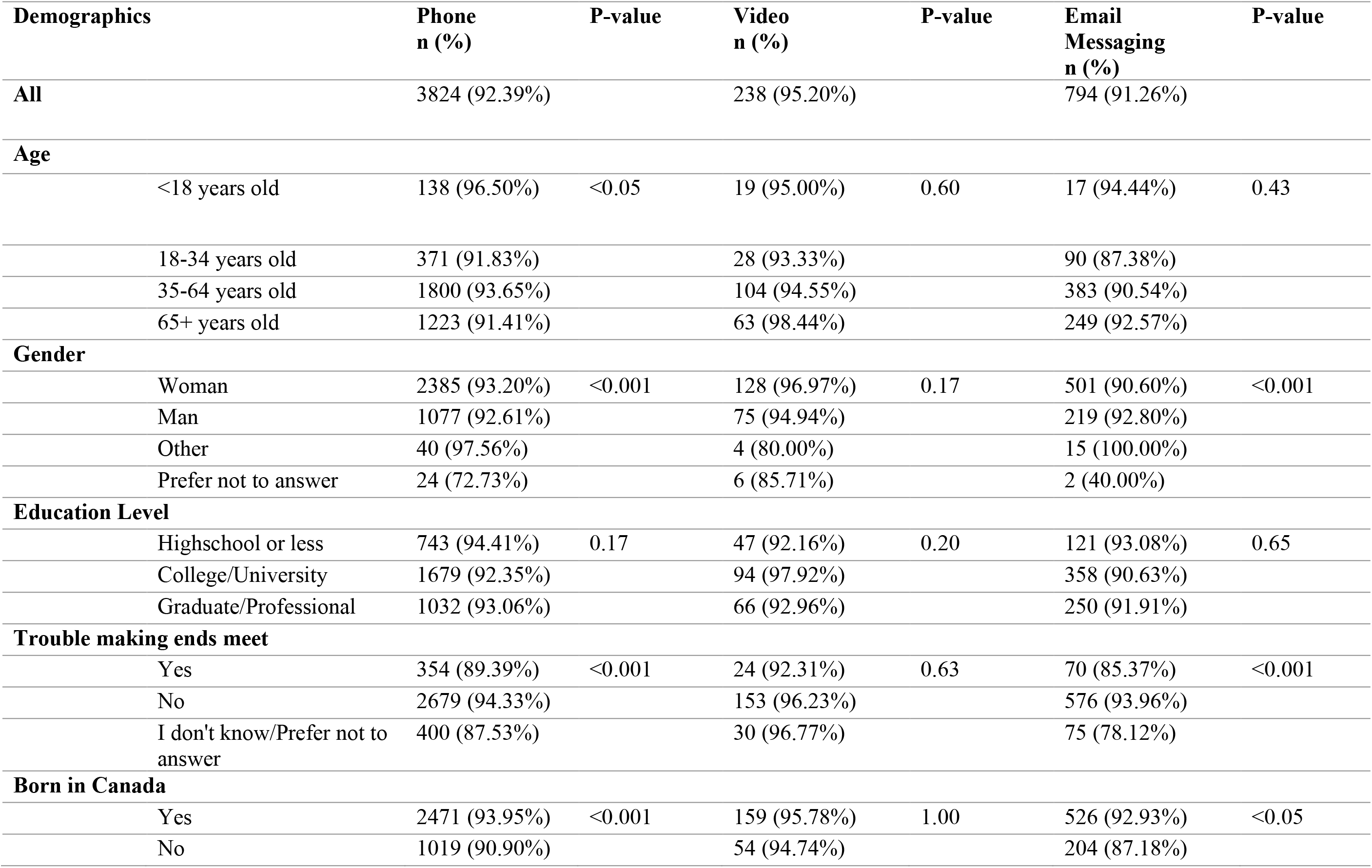
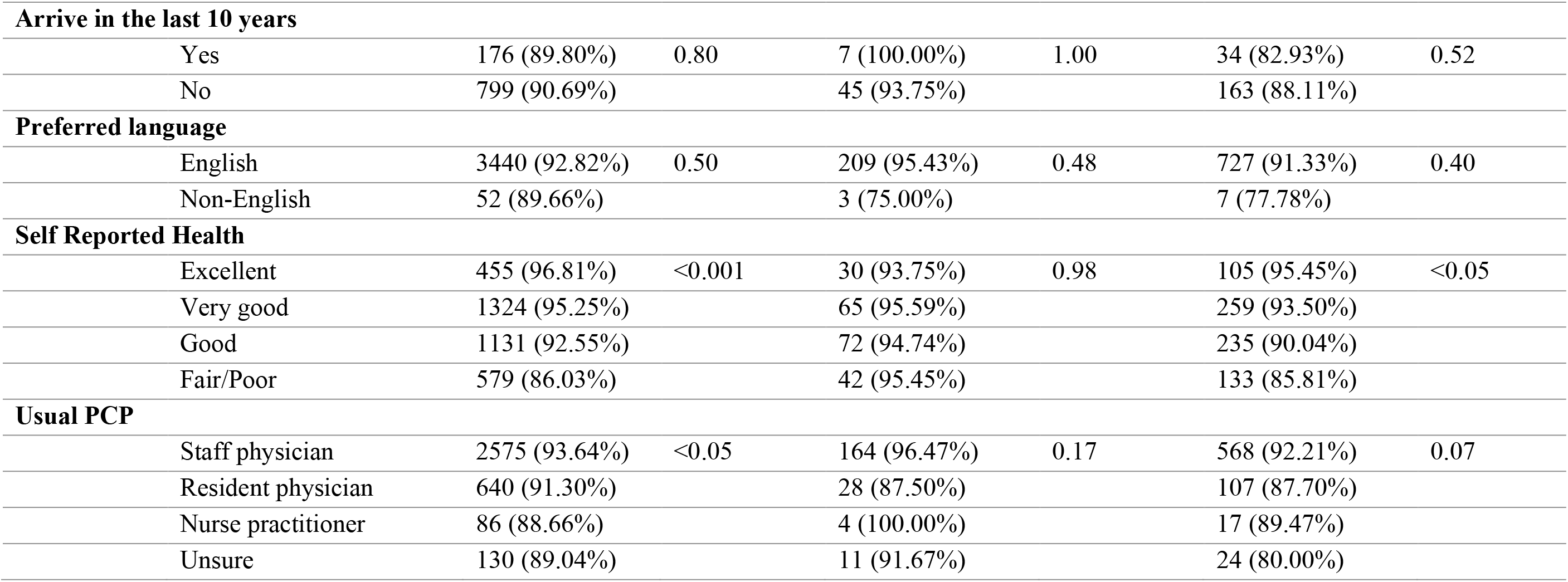
Percentage of patients who reported they were comfortable with the privacy and security of using phone, video and email or secure messaging to receive care during the pandemic, by sociodemographic characteristic

| **Demographics** | **Phone**<br>**n (%)** | **P-value** | **Video**<br>**n (%)** | **P-value** | **Email**<br>**Messaging**<br>**n (%)** | **P-value** |
| --- | --- | --- | --- | --- | --- | --- |
| **All** | 3824 (92.39%) |  | 238 (95.20%) |  | 794 (91.26%) |  |
| **Age** |  |  |  |  |  |  |
| <18 years old | 138 (96.50%) | <0.05 | 19 (95.00%) | 0.60 | 17 (94.44%) | 0.43 |
| 18-34 years old | 371 (91.83%) |  | 28 (93.33%) |  | 90 (87.38%) |  |
| 35-64 years old | 1800 (93.65%) |  | 104 (94.55%) |  | 383 (90.54%) |  |
| 65+ years old | 1223 (91.41%) |  | 63 (98.44%) |  | 249 (92.57%) |  |
| **Gender** |  |  |  |  |  |  |
| Woman | 2385 (93.20%) | <0.001 | 128 (96.97%) | 0.17 | 501 (90.60%) | <0.001 |
| Man | 1077 (92.61%) |  | 75 (94.94%) |  | 219 (92.80%) |  |
| Other | 40 (97.56%) |  | 4 (80.00%) |  | 15 (100.00%) |  |
| Prefer not to answer | 24 (72.73%) |  | 6 (85.71%) |  | 2 (40.00%) |  |
| **Education Level** |  |  |  |  |  |  |
| Highschool or less | 743 (94.41%) | 0.17 | 47 (92.16%) | 0.20 | 121 (93.08%) | 0.65 |
| College/University | 1679 (92.35%) |  | 94 (97.92%) |  | 358 (90.63%) |  |
| Graduate/Professional | 1032 (93.06%) |  | 66 (92.96%) |  | 250 (91.91%) |  |
| **Trouble making ends meet** |  |  |  |  |  |  |
| Yes | 354 (89.39%) | <0.001 | 24 (92.31%) | 0.63 | 70 (85.37%) | <0.001 |
| No | 2679 (94.33%) |  | 153 (96.23%) |  | 576 (93.96%) |  |
| I don't know/Prefer not to answer | 400 (87.53%) |  | 30 (96.77%) |  | 75 (78.12%) |  |
| **Born in Canada** |  |  |  |  |  |  |
| Yes | 2471 (93.95%) | <0.001 | 159 (95.78%) | 1.00 | 526 (92.93%) | <0.05 |
| No | 1019 (90.90%) |  | 54 (94.74%) |  | 204 (87.18%) |  |
| **Arrive in the last 10 years** |  |  |  |  |  |  |
| Yes | 176 (89.80%) | 0.80 | 7 (100.00%) | 1.00 | 34 (82.93%) | 0.52 |
| No | 799 (90.69%) |  | 45 (93.75%) |  | 163 (88.11%) |  |
| **Preferred language** |  |  |  |  |  |  |
| English | 3440 (92.82%) | 0.50 | 209 (95.43%) | 0.48 | 727 (91.33%) | 0.40 |
| Non-English | 52 (89.66%) |  | 3 (75.00%) |  | 7 (77.78%) |  |
| **Self Reported Health** |  |  |  |  |  |  |
| Excellent | 455 (96.81%) | <0.001 | 30 (93.75%) | 0.98 | 105 (95.45%) | <0.05 |
| Very good | 1324 (95.25%) |  | 65 (95.59%) |  | 259 (93.50%) |  |
| Good | 1131 (92.55%) |  | 72 (94.74%) |  | 235 (90.04%) |  |
| Fair/Poor | 579 (86.03%) |  | 42 (95.45%) |  | 133 (85.81%) |  |
| **Usual PCP** |  |  |  |  |  |  |
| Staff physician | 2575 (93.64%) | <0.05 | 164 (96.47%) | 0.17 | 568 (92.21%) | 0.07 |
| Resident physician | 640 (91.30%) |  | 28 (87.50%) |  | 107 (87.70%) |  |
| Nurse practitioner | 86 (88.66%) |  | 4 (100.00%) |  | 17 (89.47%) |  |
| Unsure | 130 (89.04%) |  | 11 (91.67%) |  | 24 (80.00%) |  |

### Future preferences for virtual care

Seventy-five percent (3798/5068), 52.2% (2644/5068), and 42.9% (2172/5068) of respondents said they wanted their practice to continue offering phone, email/secure messaging, and video after the pandemic, respectively. Age, education status, financial status, immigration status and self-reported health status were significantly associated with differences in wanting ongoing use of each of the three virtual care modalities (p<0.05 for all); those over age 65, those whose education was high school or less, those reporting yes or “I don’t know” when asked about difficulty making ends meet, those born outside Canada, and those in fair or poor health reported the lowest desire for the three virtual care modalities to continue after the pandemic compared to other groups (see Table 5).

**Table 5.** Preferences for ongoing uses of virtual care options after the pandemic, by sociodemographic characteristic.

| **Demographics** | **Phone**<br>**n (%)** | **P-value** | **Video**<br>**n (%)** | **P-value** | **Email**<br>**Messaging**<br>**n (%)** | **P-value** |
| --- | --- | --- | --- | --- | --- | --- |
| **All** | 3798 (74.94%) |  | 2172 (42.86%) |  | 2644 (52.17%) |  |
| **Age** |  |  |  |  |  |  |
| <18 years old | 171 (83.82%) | <0.05 | 120 (58.82%) | <0.001 | 117 (57.35%) | <0.001 |
| 18-34 years old | 385 (85.37%) |  | 252 (55.88%) |  | 292 (64.75%) |  |
| 35-64 years old | 1886 (84.23%) |  | 1206 (53.86%) |  | 1352 (60.38%) |  |
| 65+ years old | 1291 (80.79%) |  | 549 (34.36%) |  | 834 (52.19%) |  |
| **Gender** |  |  |  |  |  |  |
| Woman | 2510 (85.23%) | <0.001 | 1418 (48.15%) | 0.45 | 1758 (59.69%) | <0.01 |
| Man | 1142 (79.20%) |  | 662 (45.91%) |  | 780 (54.09%) |  |
| Other | 37 (78.72%) |  | 23 (48.94%) |  | 31 (65.96%) |  |
| Prefer not to answer | 29 (74.36%) |  | 21 (53.85%) |  | 20 (51.28%) |  |
| **Education Level** |  |  |  |  |  |  |
| Highschool or less | 770 (80.04%) | <0.05 | 384 (39.92%) | <0.001 | 468 (48.65%) | <0.001 |
| College/University | 1777 (83.94%) |  | 993 (46.91%) |  | 1227 (57.96%) |  |
| Graduate/Professional | 1098 (84.14%) |  | 705 (54.02%) |  | 846 (64.83%) |  |
| **Trouble making ends meet** |  |  |  |  |  |  |
| Yes | 362 (82.09%) | <0.001 | 195 (44.22%) | <0.001 | 257 (58.28%) | <0.001 |
| No | 2869 (84.48%) |  | 1665 (49.03%) |  | 2037 (59.98%) |  |
| I don't know/Prefer not to answer | 387 (74.42%) |  | 204 (39.23%) |  | 232 (44.62%) |  |
| **Born in Canada** |  |  |  |  |  |  |
| Yes | 2687 (86.01%) | <0.001 | 1579 (50.54%) | <0.001 | 1890 (60.50%) | <0.001 |
| No | 1005 (77.13%) |  | 527 (40.45%) |  | 683 (52.42%) |  |
| **Arrive in the last 10 years** |  |  |  |  |  |  |
| Yes | 166 (73.78%) | 0.13 | 89 (39.56%) | 0.90 | 101 (44.89%) | <0.05 |
| No | 806 (78.63%) |  | 413 (40.29%) |  | 554 (54.05%) |  |
| **Preferred language** |  |  |  |  |  |  |
| English | 3642 (83.21%) | 0.12 | 2086 (47.66%) | 0.08 | 2532 (57.85%) | <0.05 |
| Non-English | 52 (75.36%) |  | 25 (36.23%) |  | 29 (42.03%) |  |
| **Self Reported Health** |  |  |  |  |  |  |
| Excellent | 519 (86.64%) | <0.01 | 330 (55.09%) | <0.001 | 382 (63.77%) | <0.001 |
| Very good | 1392 (84.52%) |  | 843 (51.18%) |  | 977 (59.32%) |  |
| Good | 1179 (82.22%) |  | 635 (44.28%) |  | 809 (56.42%) |  |
| Fair/Poor | 597 (79.39%) |  | 294 (39.10%) |  | 394 (52.39%) |  |
| **Usual PCP** |  |  |  |  |  |  |
| Staff physician | 2730 (84.29%) | <0.001 | 1613 (49.80%) | <0.001 | 1943 (59.99%) | <0.001 |
| Resident physician | 659 (79.98%) |  | 336 (40.78%) |  | 415 (50.36%) |  |
| Nurse practitioner | 104 (84.55%) |  | 55 (44.72%) |  | 73 (59.35%) |  |
| Unsure | 133 (74.72%) |  | 79 (44.38%) |  | 92 (51.69%) |  |

## Discussion

Our analysis of more than 7400 patient experience surveys across 13 primary care clinics during the first months of the COVID-19 pandemic found important differences in care-seeking and comfort with virtual care based on patient income, self-reported health, and other demographic characteristics. Most participants received care from their primary care clinic in some way during the study period; however, almost a third who sought care reported they delayed it due to concerns about the pandemic. Patients who had trouble making ends meet and those who reported their health as fair or poor were more likely to seek care during the pandemic, yet were also more likely to report they delayed seeking care. Patient generally reported a high degree of comfort with phone, video and email or secure messaging. But, those reporting difficulty making ends meet, poor or fair health, and arriving in Canada in the last 10 years reported lower levels of comfort and less likely to want their practice to continue offering these virtual options.

Our results, similar to other emerging literature, suggests a complex relationship between the social determinants of health and patient comfort and preference regarding accessing care through virtual tools. A US based study prior to the pandemic found that while younger patients and those with physical disabilities were more likely to use video visits to access care, those who reported lower incomes and lived in rural populations were less likely to use this modality^26^. A recent US-based primary care study found that after care shifted to a virtual-first approach during the pandemic, a significantly smaller proportion of visits overall were with people who were low income, non-white, or non-English speakers^27^. However, a Canadian based study found that similar to our participants, those with the highest care needs (older, multiple co-morbidities), were more likely to access primary care during the early months of the pandemic compared to other groups^2^.

As many predict virtual care will continue to be a part of care delivery post-pandemic, this study highlights the importance of integrating patient experience data into future care delivery planning. Similar to other recently published data ^2, 28, 29^, our experience data indicates phone was by far the most utilized modality of virtual care and overall, participants were comfortable using virtual modalities to receive care. Patients who report financial troubles and poor health, had a higher percentage reporting accessing care (virtually and in person) during the pandemic compared to other groups; however, they reported greater concerns with the privacy and security of virtual care and less desire for virtual care to be an ongoing part of their primary care experience. This suggests that while public health measures may have pushed populations with the highest care needs to use virtual care, these modalities did not provide all patients with an equitable, patient-centered care experience. Further research should explore reasons behind the relative discomfort and low interest in virtual care and how barriers could be addressed. While access to technology may be part of this problem, other factors such as health and digital literacy, and support from peers and health care providers may also be significant^30^. Without further patient experience and demographic data to understand the ongoing use of virtual care, we risk leaving behind those who need care most.

### Strengths and Limitations

Our study had several key strengths and limitations. Our study included a large sample of respondents from multiple clinics across both urban and suburban communities. Patients were randomly sampled using birth month. Survey questions were relevant to COVID-19 and informed by primary care leaders and patients. However, our findings are open to selection bias because of the response rate, mode of delivery, and the survey being offered primarily in English; however, demographics of our sample confirm that we reached a diverse group of patients. We found substantial differences in utilization and perspectives of virtual care by sociodemographic characteristics, but these may be an underestimate of true differences. Our survey reports on experience during the early phase of the pandemic and patients’ comfort and preferences may have evolved since. Finally, although our sample was taken from 13 primary care practices, these were all academic practices in the Greater Toronto Area where physicians were paid by capitation which may limit the generalizability of the findings.

## Conclusions

We found that most patients were comfortable using virtual modalities and wanted virtual care options to continue. However, there were important differences by sociodemographic characteristics, for example, with those having difficulty making ends meet, reporting poor or fair health, and being born outside of Canada being less likely to report comfort with virtual modalities and less likely to want virtual care options to continue post-pandemic. Moving forward, clinicians and system decision makers need to carefully consider how we integrate virtual care into practices to ensure equity in access to primary care.

## Supporting information

Patient Experience Survey

## Data Availability

Following peer-reviewed publication, de-identified data can be made available on request for proposed projects advancing the public good. Please contact.

## Conflicts of interest

The authors declare no conflicts of interest.

### Prior presentations

None

## Author contributions

PA and TK conceived of and designed the study together. RW and CM conducted the analysis. All authors helped interpret the data. PA drafted the manuscript with the support of TK and all authors critically reviewed it. All authors read and approved the final manuscript

## Acknowledgements

We wish to thank Ali Damji, Debbie Elman, Frances Cousins, Jennifer Stulberg, Joanne Laine- Gossin, Karuna Gupta, Linda Weber, Melissa Witty, Navsheer Gill Toor, Noor Ramji, Sakina Walji, Sam Tirkos, Susanna Fung, Susie Kim, Thuy Nga Pham, Tiffany Florindo for their support in developing and implementing the survey at their respective teaching practices, Trish O’Brien and Kirsten Eldridge for their support with survey implementation across all sites, Danielle Martin for her feedback on our draft manuscript, and the patient partners who helped us refine the survey questions.

## Appendix A: Site Demographics

| Site name | City | Number of surveys sent<br>(N= 32, 307) | Number of responses<br>(N=7,532) | Response Rate<br>(%) |
| --- | --- | --- | --- | --- |
| **Barrie and Community Family Health Team** | Barrie | 748 | 279 | 37% |
| **Credit Valley Family Health Team** | Mississauga | 2,056 | 645 | 31% |
| **Health for All Family Health Team** | Markham | 1,900 | 535 | 28% |
| **Mount Sinai Academic Family Health Team** | Toronto | 2,100 | 391 | 19% |
| **Southlake Family Health Team** | Newmarket | 1,968 | 735 | 37% |
| **North York Family Health Team** | North York | 1,466 | 390 | 27% |
| **Platinum Medical Family Health Organization** | Scarborough | 1,792 | 222 | 12% |
| **Sunnybrook Academic Family Health Team** | Toronto | 1,447 | 409 | 28% |
| **St. Joseph's Health Centre Family Medicine/Urban Family Health Team** | Toronto | 1,135 | 265 | 23% |
| **St. Michael's Hospital Academic Family Health Team** | Toronto | 5,180 | 1337 | 26% |
| **South East Toronto Family Health Team** | Toronto | 5,244 | 591 | 11% |
| **Summerville Family Health Team** | Mississauga | 5,070 | 1426 | 28% |
| **Women's College Hospital Family Practice Health Centre** | Toronto | 2,201 | 307 | 14% |

