## Supplementary material for "Sociodemographic differences in patient experience with virtual care during COVID-19": Patient Experience Survey

### Improving your Patient Experience

Dept of Family and Community Medicine, University of Toronto
Draft Jun 1, 2020

Start of Block: Intro Block

Q1 Dear THE CLINIC'S NAME Patient,

The CLINIC wants to know about your experience getting health care during the COVID-19 pandemic. We are asking you to complete a short survey, which will take about 5 minutes. Your answers will help us to improve the care we provide.

You are receiving this survey because either you or your family member is a patient with CLINIC and have a birthday in MONTH1, MONTH2, MONTH3. Participation is voluntary and responses are confidential. We do not ask for your name in the survey and your answers cannot be linked back to your chart. We are interested in your honest opinion, whether it is negative or positive. Your responses to this survey will not change the care you receive from us.

PLEASE NOTE: This survey is for the person in your family who has a birthday in MONTH1, MONTH2, MONTH3. If this person is someone you are a caregiver for (a child or parent), please respond based on their care experience. If your own birthday is also in MONTH 1, MONTH 2, MONTH 3, you can choose to respond based on your own care experience.

End of Block: Intro Block

Start of Block: Block 1

Q2 Section 1 – Care needs during pandemic
*The following questions will help us better understand your comfort with accessing care during the COVID-19 pandemic. Please think about the care you received after March 17, 2020 (the date Ontario declared a state of emergency due to COVID-19).*

Q3 **Did you receive care from a doctor, nurse or healthcare provider at CLINIC during the COVID-19 pandemic? This includes care delivered in person, by phone, by video or by email or secure message.**

- ◯ Yes (1)
- ◯ No (2)

*Skip To: Q12 If DFCM1.1 = No*

Q4 **How did you receive care during this time?** (Select all that apply)

- ▢ In person (1)
- ▢ Phone call (2)
- ▢ Video (3)
- ▢ Email or secure message (4)

*Display This Question:*

*If DFCM1.2 = Phone call*

Q5 When using the PHONE to discuss your health concerns, how comfortable were you with the level of privacy and security? (Select one response)

- Extremely comfortable (1)
- Somewhat comfortable (2)
- Neither comfortable nor uncomfortable (3)
- Somewhat uncomfortable (4)
- Extremely uncomfortable (5)

---

*Display This Question:*

*If DFCM1.2 = Phone call*

X→

Q6 **When using the PHONE , was there anything you did not talk about because you were worried about privacy?** (Select one response with optional comments)

- Yes (1)
- No (2)
- Comments: (3) ________________________________________________

---

*Display This Question:*

*If DFCM1.2 = Video*

X→

Q7 When using VIDEO to discuss your health concerns, how comfortable were you with the level of privacy and security?  (Select one response)

- ○ Extremely comfortable  (1)
- ○ Somewhat comfortable  (2)
- ○ Neither comfortable nor uncomfortable  (3)
- ○ Somewhat uncomfortable  (4)
- ○ Extremely uncomfortable  (5)

---

*Display This Question:*

*If DFCM1.2 = Video*

X→

Q8 **When using VIDEO, was there anything you did not talk about because you were worried about privacy?** (Select one response with optional comments)

- ☐ Yes  (1)
- ☐ No  (2)
- ☐ Comments:  (3) ________________________________________________

---

*Display This Question:*

*If DFCM1.2 = Email or secure message*

X→

Q9 When using EMAIL or SECURE MESSAGE to discuss your health concerns, how comfortable were you with the level of privacy and security? (Select one response)

- Extremely comfortable (1)
- Somewhat comfortable (2)
- Neither comfortable nor uncomfortable (3)
- Somewhat uncomfortable (4)
- Extremely uncomfortable (5)

---

*Display This Question:*

*If DFCM1.2 = Email or secure message*

X→

Q10 **When using EMAIL or SECURE MESSAGE, was there anything you did not talk about because you were worried about privacy?** (Select one response with optional comments)

- Yes (1)
- No (2)
- Comments: (3) ________________________________________________

---

X→

Q11 **Did you avoid or delay receiving care from THE CLINIC because of the COVID-19 pandemic?** (Select one response)

- Yes (1)
- No (2)

---

*Display This Question:*

*If DFCM1.1 = No*

Q12 **Please tell us why you did not get care from THE CLINIC during the COVID pandemic:** (Select all that apply)

- ▢ I did not have any health needs (1)
- ▢ I did not know I could receive care from the clinic during the pandemic (2)
- ▢ I did not want to come into the clinic because I was worried for my personal safety (3)
- ▢ I tried but could not get an appointment (4)
- ▢ The hours were inconvenient (5)
- ▢ I could not get through to the clinic on the phone (7)
- ▢ Other (please specify): (6) ________________________________________________

*Display This Question:*

*If DFCM1.1 = No*

Q13 **Did you get care somewhere else during the COVID pandemic?** (Select all that apply)

- [x] No, I did not get care elsewhere (1)
- [ ] I got care from a walk-in clinic in person (2)
- [ ] I got care from a walk-in clinic by phone or video (3)
- [ ] I went to the emergency department (4)
- [ ] Other (please specify): (5) ________________________________________________

End of Block: Block 1

---

Start of Block: Block 2

Q14 **Section 2: Getting URGENT CARE when you are sick during the COVID-19 pandemic**
*The following questions help us better understand the experience of patients who were sick and wanted to be seen urgently. Please answer the questions below for the time during the COVID-19 pandemic only (starting March 17, 2020).*

---

Q15 **During the COVID-19 pandemic, was there a time when you were sick and URGENTLY needed care at THE CLINC?** (Select one response)

- ○ Yes (1)
- ○ No (2)

*Skip To: End of Block If DFCM2.1 = No*

---

Q16 **Think about the time you needed URGENT CARE. How many days did it take from when you first tried to book an appointment at our clinic to when you received care?**

**Care could include an in-person visit, phone visit, video visit and/or email or secure messaging.** (Select one response).

- ◯ On the same day (1)
- ◯ The next day (2)
- ◯ In 2 to 3 days (3)
- ◯ In 4 to 7 days (4)
- ◯ After more than 1 week (5)
- ◯ Never able to get an appointment (6)
- ◯ Not sure (7)

---

*Display This Question:*

*If DFCM2.2 = In 2 to 3 days*

*Or DFCM2.2 = In 4 to 7 days*

*And DFCM2.2 = After more than 1 week*

*And DFCM2.2 = Never able to get an appointment*

Q17 **Why were you not able to get care the same or next day?** (Select all that apply)

- ☐ I was informed that there was no availability (1)
- ☐ I was offered an appointment but not with the provider I preferred (2)
- ☐ I was offered an appointment but not at the time I preferred (3)
- ☐ I could not get through to the clinic on the phone (4)
- ☐ Other (Please specify) (5) ________________________________________________

---

Q18 How would you describe the length of time it took between making the appointment and receiving care? (Select one response)

- ◯ About right (1)
- ◯ Somewhat too long (2)
- ◯ Much too long (3)

---

Q19 During the COVID-19 pandemic, did you need urgent care on an evening, weekend, or public holiday?

- ◯ Yes (1)
- ◯ No (2)

---

*Display This Question:*

*If DFCM2.5 = Yes*

Q20 How easy or difficult was it to get urgent care from CLINIC on an evening, weekend, or holiday during the COVID-19 pandemic?

- ◯ Very easy (1)
- ◯ Somewhat easy (2)
- ◯ Neither easy nor difficult (3)
- ◯ Somewhat difficult (4)
- ◯ Very difficult (5)

**End of Block: Block 2**

---

**Start of Block: Block 3**

Q21 ***Section 3: Care experience***
*For the next set of questions, please think about your experience when receiving care from your doctor or nurse practitioner during the COVID-19 pandemic only (starting March 17, 2020). This includes care delivered in person, by phone, by video or by email or secure message.*

---

Q22 **How often did you receive care from the doctor or nurse practitioner that you prefer?** (select one response)

- ○ I do not have a preferred health care provider (1)
- ○ Always (2)
- ○ Usually (3)
- ○ Occasionally (4)
- ○ Rarely (5)
- ○ Never (6)

---

Q23 **How often did you receive care within a reasonable time from your doctor or nurse practitioner**? (Select one response)

- ○ Always (1)
- ○ Usually (2)
- ○ Occasionally (3)
- ○ Rarely (4)
- ○ Never (5)

---

Page Break

Q24 **When you received care from your doctor or nurse practitioner, how often did they involve you as much as you want to be in decisions about your care and treatment?** (Select one response)

- Always (1)
- Usually (2)
- Occasionally (3)
- Rarely (4)
- Never (5)

---

Q25 **When you received care from your doctor or nurse practitioner, how often did they spend enough time with you?** (Select one response)

- Always (1)
- Usually (2)
- Occasionally (3)
- Rarely (4)
- Never (5)

End of Block: Block 3

---

Start of Block: Block ADDITIONAL

Q26 Click to write the question text

End of Block: Block ADDITIONAL

---

Start of Block: Block 4

Q27 **Section 4: Your recommendations** *For the next set of questions, please share your thoughts on how we can improve THE CLINIC.*

---

Q28 **After the COVID-19 pandemic is over, which of these care options should the clinic continue to offer?** (Select all that apply)

- ▢ Phone (1)
- ▢ Video (2)
- ▢ Email/secure messaging portal (3)
- ▢ Other (4) ________________________________________________
- ▢ None of the above (5)

---

Q29 **What changes did our clinic make during COVID-19 that you would like us to continue?** (Optional)

________________________________________________________________

________________________________________________________________

________________________________________________________________

________________________________________________________________

________________________________________________________________

---

Q30 **What do you think our clinic could have done differently to better meet your health needs during the COVID-19 pandemic?** (Optional)

________________________________________________________________

________________________________________________________________

Q31 **Overall, would you recommend our clinic, to your friends and family?** (Select one response)

- ◯ Yes (1)
- ◯ No (2)

End of Block: Block 4

Start of Block: Block 5a

Q32 **Are you filling this survey out on behalf of someone else?** (Select one response)

- ◯ Yes (1)
- ◯ No (2)

End of Block: Block 5a

Start of Block: Block 5b - Family

Q33 **Section 5: About Your Family Member**
This final section of the survey helps us understand if some groups are experiencing care differently than others.

Q34 I am filling this survey on behalf of my child or family member who is: (optional)

- ○ 0-5 years old (1)
- ○ 6-17 years old (2)
- ○ 18-24 years old (3)
- ○ 25-34 years old (4)
- ○ 35-49 years old (5)
- ○ 50-64 years old (6)
- ○ 65-79 years old (7)
- ○ 80+ years old (8)

Q35 What gender do they identify with? (optional)

- ○ Woman/girl (1)
- ○ Man/boy (2)
- ○ Transgender woman/girl (3)
- ○ Transgender man/boy (4)
- ○ Non-binary (for example gender queer, 2-spirit) (5)
- ○ Identity not listed (please specify) (6) ________________________________________________
- ○ Prefer not to answer (7)

Q36 What is their highest level of education? (optional)

- ◯ Elementary school or less (1)
- ◯ Some High school (2)
- ◯ High School Diploma (3)
- ◯ College or University Diploma Degree (4)
- ◯ Graduate or Professional Degree (5)

---

Q37 **Do they have trouble making ends meet (money problems) at the end of the month?** (optional)

- ◯ Yes (1)
- ◯ No (2)
- ◯ I don't know (3)
- ◯ Prefer not to answer (4)

---

Q38 Were they born in Canada? (optional)

- ◯ Yes (1)
- ◯ No (2)

---

*Display This Question:*

*If DFCM5b.5 = No*

Q39 **Did you arrive in Canada in the last 10 years?** (optional)

- ○ Yes (1)
- ○ No (2)

---

Q40 **What language would they prefer speaking with their primary care provider?** (optional)

- ○ English (1)
- ○ French (2)
- ○ Other (please specify): (3) ________________________________________________

---

Q41 In general, would you say their health is: (optional)

- ○ Excellent (1)
- ○ Very Good (2)
- ○ Good (3)
- ○ Fair (4)
- ○ Poor (5)

---

Q42 What is their postal code? (optional)

________________________________________________________________

---

Q43 Which primary care provider do they usually see? (optional)

- Staff Physician (1)
- Resident Physician (2)
- Nurse Practitioner (3)
- Unsure (4)

End of Block: Block 5b - Family

---

Start of Block: Block 5c - Yourself

Q44 **Section 5: About You**
This final section of the survey helps us understand if some groups are experiencing care differently than others.

Q45 **How old are you?** (optional)

- 0-5 years old (1)
- 6-17 years old (2)
- 18-24 years old (3)
- 25-34 years old (4)
- 35-49 years old (5)
- 50-64 years old (6)
- 65-79 years old (7)
- 80+ years old (8)

Q46 **What gender do you identify with?** (optional)

- ○ Woman/girl (1)
- ○ Man/boy (2)
- ○ Transgender woman/girl (3)
- ○ Transgender man/boy (4)
- ○ Non-binary (for example gender queer, 2-spirit) (5)
- ○ Identity not listed (please specify) (6) ________________________________________________
- ○ Prefer not to answer (7)

Q47 **What is your highest level of education?** (optional)

- ○ Elementary school or less (1)
- ○ Some High school (2)
- ○ High School Diploma (3)
- ○ College or University Diploma Degree (4)
- ○ Graduate or Professional Degree (5)

Q48 **Do you have trouble making ends meet (money problems) at the end of the month?** (optional)

- Yes (1)
- No (2)
- I don't know (3)
- Prefer not to answer (4)

---

Q49 **Were you born in Canada?** (optional)

- Yes (1)
- No (2)

---

*Display This Question:*

*If DFCM5c.5 = No*

Q50 **Did you arrive in Canada in the last 10 years?** (optional)

- Yes (1)
- No (2)

---

Q51 **What language would you prefer speaking with your primary care provider?** (optional)

- ○ English (1)
- ○ French (2)
- ○ Other (please specify): (3) ________________________________________________

Q52 **In general, would you say your health is:** (optional)

- ○ Excellent (1)
- ○ Very Good (2)
- ○ Good (3)
- ○ Fair (4)
- ○ Poor (5)

Q53 What is your postal code? (optional)

________________________________________________________________

Q54 Which primary care provider do you usually see? (optional)

- Staff Physician (1)
- Resident Physician (2)
- Nurse Practitioner (3)
- Unsure (4)

End of Block: Block 5c - Yourself

---

Start of Block: End of Survey

Q55 Thank you for spending the time to complete this survey. Note: Please click "Submit" to record all your answers.

End of Block: End of Survey

---
